# Comparison of the capacity of several machine learning tools to assist immunofluorescence-based detection of anti-neutrophil cytoplasmic antibodies

**DOI:** 10.1101/2024.01.26.24301725

**Authors:** Daniel Bertin, Pierre Bongrand, Nathalie Bardin

**Affiliations:** Service d’Immunologie, Biogénopôle, Hôpital de la Timone, Assistance Publique-Hôpitaux de Marseille (AP-HM), Marseille, France; Aix-Marseille Univ, Laboratoire Adhésion et Inflammation, UM61, Marseille, France; INSERM U1067, France; CNRS, U7333, France; INSERM, U1076, France; Aix Marseille Univ, INSERM, INRAE, C2VN, Marseille, France

**Keywords:** Artificial intelligence, ANCA, immunofluorescence, vasculitis, image analysis, myeloperoxdase, proteinase 3

## Abstract

The success of artificial intelligence and machine learning is an incentive to develop new algorithms to increase the rapidity and reliability of medical diagnosis. Here we compared different strategies aimed at processing microscope images used to detect anti-neutrophil cytoplasmic antibodies, an important vasculitis marker: (i) basic classifier methods (logistic regression, k-nearest neighbors and decision tree) were used to process custom-made indices derived from immunofluorescence images yielded by 137 sera. (ii) These methods were combined with dimensional reduction to analyze 1733 individual cell images. iii) More complex models based on neural networks were used to analyze the same dataset. The efficiency of discriminating between positive and negative samples and different fluorescence patterns was quantified with Rand-type accuracy index, kappa index and ROC curve. It is concluded that basic models trained on a limited dataset allowed positive/negative discrimination with an efficiency comparable to that obtained by conventional analysis performed by humans (0.84 kappa score). More extensive datasets may be required for efficient discrimination between different fluorescence patterns generated by different auto-antibody species.

## 1 Introduction

The steady growth of the diversity, power and cost of therapeutic tools is an incentive to attempt at increasing the precision of diagnosis without a parallel increase of expenses. The spectacular progress of computer-based methods, referred to as artificial intelligence (AI) or machine learning (ML) may be of considerable help in this respect, by allowing to extract maximal information from biological data with optimal rapidity and minimal recourse to biological experts. Despite initial disappointment met several decades ago along this line [1], the tremendous progress of machine learning algorithms resulted in a steady development of the use of AI in medicine [2], be it to process large datasets generated by multi-omic methods in order to elaborate general prediction algorithms [3], identify new markers of clinical interest [4] or to analyze the output of standard biological tests performed on individual patients in order to achieve more rapid, more reliable and less costly diagnosis [5].

Indirect immunofluorescence has long been considered as an important tool, and even a so-called gold standard, to detect anti-nuclear antibodies (ANAs) associated to severe conditions such as systemic lupus erythematosus [6][6], or anti-neutrophil cytoplasmic antibodies (ANCAs) that are associated to a number of vasculitis-involving syndromes [7]. The basic principle consists of exposing fixed cells to patients’ sera, and looking for the presence of auto-antibodies by microscopical observation of slides labeled with fluorescent anti-immunoglobulin antibodies. ANAs are usually detected on Hep-2 cells, and ANCAs on polymorphonuclear leukocytes. An experienced pathologist is required to recognize specific patterns revealing the presence of suspected antibodies. Thus, the examination of fluorescence patterns on ethanol-fixed leukocytes may reveal so-called cellular-type ANCAs (C-ANCAs) with a cytoplasmic pattern usually associated to anti-proteinase 3 antibodies, or perinuclear-type ANCAs (P-ANCAs), usually associated to anti-myeloperoxidase antibodies [8]. Other patterns may be due to ANAs or antibodies of other specificities that may be indicative of different pathological situations with different therapeutic implications [9] [10]. The well recognized finding [11] [12] [13] that inconsistencies may occur between different laboratories is a strong incentive to attempt as standardizing the processing of immunofluorescence images [14]. While numerous attempts have long been made at developing automatic diagnostic procedures, these methods required the definition, measurement and processing of numerous so-called hand-crafted [11] texture parameters [15]. It was thus an attractive prospect to take advantage of rapidly developing ML methods that allow to build classification algorithms by autonomous treatment of training datasets and met with impressive success in important domains such as text or facial recognition [16] [17]. Further, the availability of these algorithms is strongly increased by the development of open access platforms such as scikit learn (http://scikit-learn.org) or tensor flow(https://www.tensorflow.org/), that include exhaustive online documentation and the use of which is facilitated by excellent written tutorials [18] [19]. Accordingly, these platforms are currently used in state-of-the-art research projects [20] [21].

An essential requirement to foster progress is to make use of objective tools for measuring the efficiency of different classification methods. The fraction of accurate predictions, that may be designated as predictive accuracy (*pa*), is a widely used and fairly intuitive reporter of the efficiency of binary classification [11] However, it may provide a less appropriate measure of the efficiency of multiclass data partition, that may be more precisely represented by parameters such as Rand index [22]. Also, a calculated classification accuracy may be deceptive. Indeed, if an algorithm is used to detect positive samples in a batch of sera that are mostly negative - a quite common situation - a very high accuracy may be obtained by classifying all samples as negative ! A widely used correction [23] consists of calculating the accuracy increase provided by a model as compared to random agreement following a simple equation, yielding so-called Cohen *kappa index*.

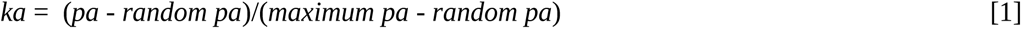

where *ka* is for kappa index, *random pa* is he precision accuracy corresponding to a random choice, and *maximum pa* the precision provided by a fully exact model. This index was used in numerous reports of diagnostic accuracy [24] [25] [12]. It was suggested to consider the agreement as moderate, substantial or perfect when kappa index is respectively higher than 0.4, 0.6 and 0.8 [23]. It must be kept in mind that this index is dependent on diagnostic criteria as well as the specific features of the sample population used to perform the comparison between a given model and a gold standard. It may thus be appropriate to mention “kappa-type measures” [23]and give detail on the exact algorithm used to calculate kappa index. Further, a more exhaustive account of the efficiency of a method is provided by plotting *sensitivity* (i.e. the fraction of positive samples that are classified as positive) versus *one minus specificity* (where specificity is the fraction of negative samples that are classified as negative). This is called the receiver-operator-characteristic (ROC) curve [26], and the model efficiency is expressed as the area under the curve (*auc*) that is expected to be comprised between 0.5 (corresponding to a random classification) and 1 (corresponding to a perfect classification. It is important to recall that this curve is dependent on the population used to perform the comparison. In a recent metaanalysis of 56 reports [27], the minimal *auc* value required for a test to be considered as good or very good ranged between 0.75 and about 0.95. Also, it may be useful to use an index suited for both binary and multilabel classification. A Rand-type index corrected for random agreement elaborated by the scikit-learn tem was found convenient in a recent study [28]. This index will be designated here as corrected predictive accuracy (*cpa*). As shown below, this was found to be tightyly related to *pa*, kappa index and *auc* (see section 2.1 and Figure 2).

In view of the diagnostic importance of ANA or ANCA detection, automatic methods have long been elaborated to obviate the aforementioned problems associated with visual microscopic examinations [29]. The starting point was the use of image analysis and procedures such as image segmentation and texture quantification. This allowed to develop commercially available systems that reliably performed simple tasks such as discrimination between positive and negative samples [30]. Thus, a simple algorithm elaborated in our laboratory allowed safe discrimination between ANA-positive and negative samples with a kappa coefficient of 0.92 [25] comparable to values obtained with a number of commercial systems [30]. However, the recognition of specific patterns appears more challenging with a recognition efficiency of some commercial systems varying between about 40% and 85% [30]. As recently reviewed [11], numerous reports dealt with the application of recent ML tools to the recognition of ANA patterns. However, ANCA detection may be considered as more demanding since neutrophils display more complex nucleus shapes than Hep-2 that are used for ANA detection [24] [29]. Further, a limitation of commercial systems is that is is difficult to freely improve available tools due to the complexity and limited availability of the algorithms they use.

The aim of the present report was to present a detailed description of the potential of currently available machine learning tools to classify immunofluorescence images used for ANCA detection. We first built a dataset including 1733 cell images obtained by processing 137 sera. Two strategies were followed. First, microscopic images were used to extract four features suggested by biological experience, and assess the capacity of basic machine learning tools including logistic regression, K neighbor classifier and decision tree. Secondly, cell images (2500 pixels) were subjected to individual analysis with aforementionded models and more complex neural networks. It is concluded that, while *kappa scores* higher than 0.8 were easily obtained for discrimination between positive and negative samples, more work and more extensive datasets are required to allow pattern classification. The promise and pitfalls of several development strategies are exemplified by preliminary data.

## 2 Methods

### 2.1 Patients

This retrospective study was performed on 137 sera processed in the immunology laboratory of Marseilles public hospitals for detection of antineutrophil cytoplasmic antibody (ANCAs), as requested by clinical departements. All serum samples were part of Marseilles Biobank (registered as DC 2012_1704) and the study was approved by the medical evaluation board and health data committee of Assistance Publique-Hôpitaux de Marseille, Marseille, France and fulfilled local requirements in terms of data collection and protection of data (GDPR 2019-133).

### 2.2 Immunofluorescence

ANCA and their staining pattern (perinuclear, cytoplasmic) were detected by IF on ethanol-fixed human neutrophil slides (Immuno Concepts, CA, USA) [31]. Serum samples diluted in phosphate-buffered saline were added for 30 minutes at room temperature (RT). After washing, bound antibodies were labeled by incubation with fluorescein isothiocyanate (FITC)-conjugated sheep anti-human immunoglobulin (Immuno Concepts, CA, USA) for 30 minutes at RT. Slides were then washed and embededded with a 4,6-diaminophenylindol (DAPI) containing medium (Vectashield, Vector laboratories inc, Burlingame, Ca) for nuclear staining.

For each patient, two images of a same central microscopic field were automatically captured with 20x objective at two different excitation wavelengths: 480 nm for FITC stain and 360 nm for DAPI stain. We used a fully robotized fluorescence microscope (Axio Imager M2, Carl Zeiss, Jena, Germany) equipped with an automated 200-slide handling system (SlideExpress, Märzhäuser, Wetzlar, Germany) and with 360 nm and 480 nm LEDs for excitation (Colibri 2 LED illumination system, Carl Zeiss, Jena Germany). Images with 1360 × 1024 pixels resolution were captured using a monochrome CCD camera (ProgRes® MF Cool camera, Jenoptik, Germany) with pixel size of 6.45 μm ^2^ . Exposure times for FITC and DAPI captures were 70 ms and 200 ms respectively. All captured grayscale images were 8 bit-depth and have been saved in Tagged Image File Format (TIFF) as previously described [14]. Representative images are shown on Figure 1.

**Figure 1.**
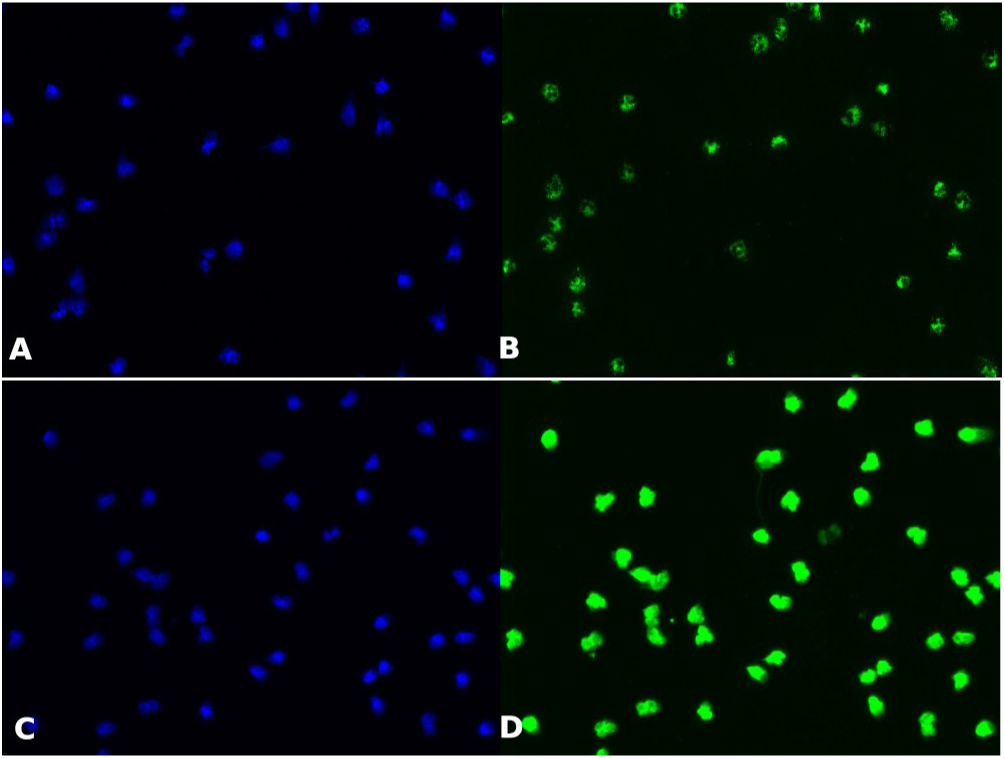
Representative Microscopic images. Ethanol fixed neutrophils were processed for immunofluorescence with a serum positive for C-ANCA (A-B) or P-ANCA (C-D) and the localization of DAPI (A-C) or FITC (C-D) was revealed by fluorescence microscopy.

### 2.3 Elisa testing

The specificity of samples classified as positive was checked by looking for anti-myeloperoxidase or anti-proteinase-3 antibodies by Enzyme-linked immunoassay (ELISA, Euroimmune, Lübeck, Germany).

### 2.4 Image processing

#### 2.4.1 calculation of overall quantitative indices

Images were first processed with a previously described software (ICARE) [25]written in java as a plugin to Image J [32], using an additional custom-made plugin allowing automatic determination of four quantitative indices that were felt relevant to the presence of ANCAs. Briefly, the DAPI image was used to define cell surface as the set of pixels with an intensity at least fourfold higher than the first peak on the intensity histogram. Unexpectedly, while this matched the nucleus on Hep-2 cells (used for ANA detection), it filled whole neutrophil surfaces, likely due to the contorted nucleus shape and possibly particular cytoplasmic staining properties or this cell population. This was used as a basis for the determination of the following four indices :

- Index i1 is the ratio between the mean intensity on FITC images of pixels classified as “inside” and “outside pixels”. This was expected to allow discrimination between positive and negative samples.
- Index i2 is the ratio between the mean FITC intensity of pixels defined as “inside” and the first peak intensity of the histogram of FITC image.
- Index i3 is similar to i2, but inside is defined on DAPI histograms as pixels with intensity 16-fold higher than that of the first background. It was expected that this region might be closer to actual nuclear regions.
- Index i4 is the correlation between FITC and DAPI pixel intensities in regions defined as “inside” on DAPI images. It might be hoped that the correlation would be highest with ANA, lowest with C-ANCA and intermediate with P-ANCA.

Images were simultaneously classified by an experienced pathologist as negative (0), C-ANCA (1), P-ANCA (2) or atypical/ANA type (3) and processed by ICARE [25]. A file including 137 samples with 4 indices each was thus prepared for ML processing. This allowed to build a dataset including 137 samples (102 negative, 9 C-ANCA, 21 P-ANCA, 5 atypical/ANA).

#### 2.4.2 Building individual cell images

Individual cell images were built out of whole microscopic fields with both following treatments:

- First, cell region was defined on DAPI images with a threshold-based algorithm that we used for decades as a software that was first written in assembly language [33], then translated into C++ [34] and finally as a java plugin for Image J [32].
- Secondly, areas limited with DAPI-determined boundaries were resized to 50×50 pixel images by plain homothety with a custom-made python program and stored as csv files for further treatment. This allowed to build a dataset of 1733 individual cell images (513 negative, 309 C-ANCA, 789 P-ANCA, 122 atypical/ANA).

### 2.5 Machine learning

#### 2.5.1 Classification based on “hand-crafted” parameters

The 137-sample csv file was processed as a four-parameter numpy array with four standard models provided by scikit-learn as previously described [28]: logistic regression classifier, k nearest neighbor classifier, decision tree classifier and multilayer perceptron as a simple neural network. Since the datasets were not extensive enough to allow hyperparameter optimization, we essentially used default values with minimal changes that were found suitable for a low feature number dataset (this was 4 as indicated above) [28]. For each method, the dataset was splitted 100 times into a training set and a test set and classification efficiency was obtained by calculating prediction accuracy (*pa*), corrected prediction accuracy (*cpa*, a modified rand type score corrected for chance, as calculated with scikit-learn adjusted_rand_score function, Cohen kappa score and area under roc score (*auc*) when positive/negative discrimination was studied.

#### 2.5.2 Analysis of individual cell images

- Individual cell images (2500 pixels) were first subjected to a scaling procedure (scikit-learn RobustScaler method) to ensure that all parameters displayed similar median and quartile distribution. In some cases, data reduction was performed with principal component analysis.
- Images were analyzed with aforementioned standard algorithms (logical regression, k-next neighbors or decision tree and neural networks). In addition to aforementioned multilayer perceptron classifier, we used convolutive networks, since they are thought to be well suited for image analysis [11] [19]. Tensorflow platform was used, taking advantage of keras application programming interface.

Under all conditions, efficiency parameters were calculated by random splitting of datasets between 10 and 100 times between a train set (about 75% of samples) and a test set (about 25% of samples). Classification efficiency was then calculated on the train and test set after training models on train sets.

As shown on Figure 2, when 32 different models and model settings were used to calculate all four indices *on the same dataset* of 1733 cells, a tight correlation was found between these indices.

**Figure 2.**
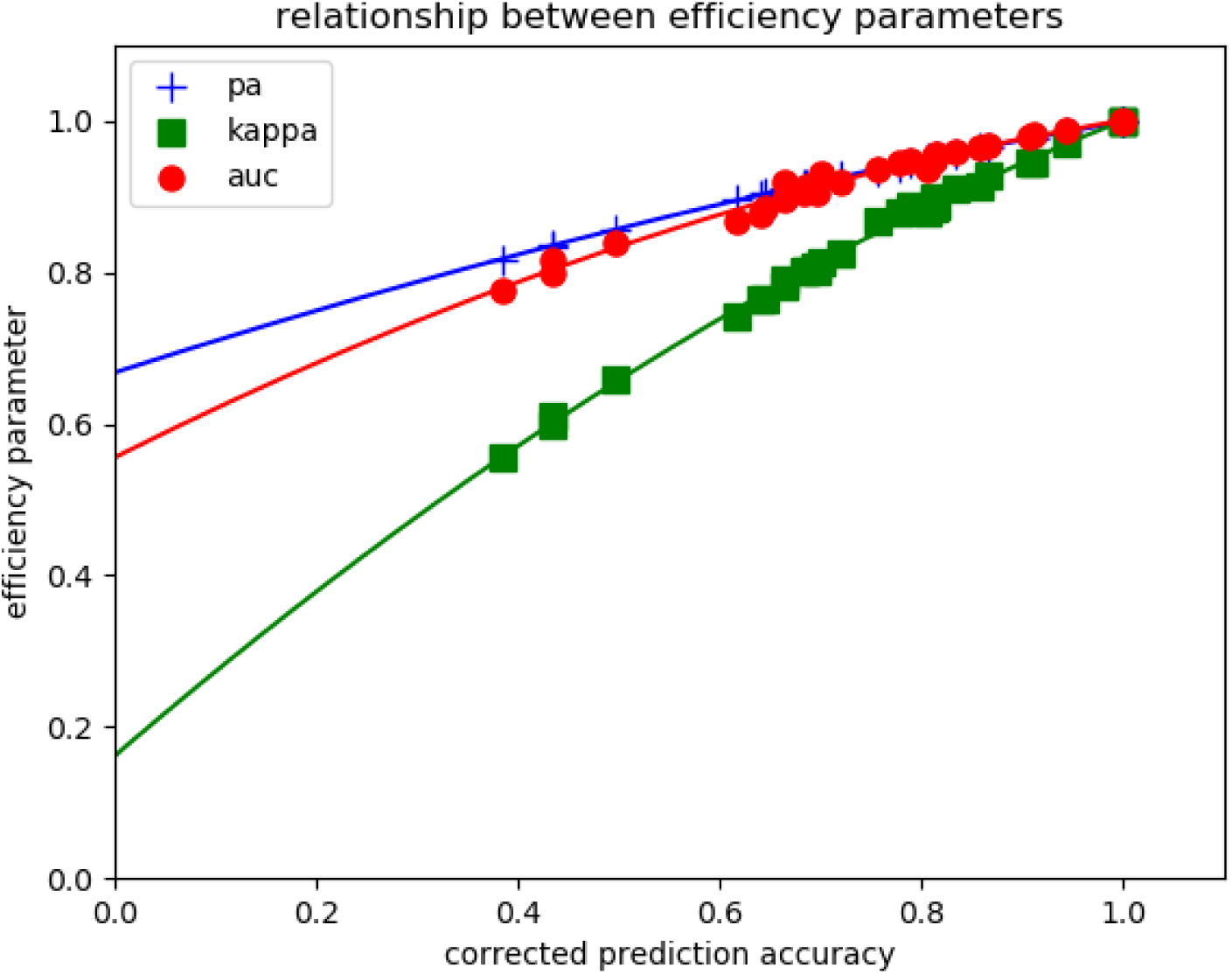
Relationship between classification efficiency indices. A series of 1733 cell images were classified with two models (k neighbors classifier and multilayer perceptron classifier) using different hyperparameters and differerent preprocessing procedures (data scaling with or without dimensional reduction based on principal component analysis). Prediction accuracy, area under roc score and Cohen-kappa score were plotted versus corrected prediction accuracy as shown.

## 3 Results

### 3.1 Combination of biologically inspired indices and machine learning

Data were used first to perform a binary classification between ANCA-positive and negative samples. Secondly, we try to discriminate between immunofluorescence patterns.

#### 3.1.1 Discrimination between positive and negative samples with full image-related indices

First, full images (encompassing entire microscopic fields) generated with individual sera were processed to derive four quantitative indices that were felt with a possible relevance to ANCAs as explained in section 2.4.1. This yielded a dataset including 137 samples that were classified as negative (102/137) or positive (35/137) by conventional observation. This dataset was randomly split 100 times into a training set (102 samples) and a test set (35 samples). As suggested by a previous comparison of the efficiency of eight standard algorithms to analyze limited datasets [28], we used three fairly simple algorithms : logistic regression (LR), k nearest neighbors (kNN) and decision trees (DT). The prediction accuracy (*pa*), corrected prediction accuracy (*cpa*) and area under roc curve (*auc*) obtained on train and test sets after training on train set are shown on Table 1.

**Table 1.** Discrimination between Anca-positive and negative sera by processing a 4-parameter dataset. 137 sera were assayed for ANCAs with indirect immunofluorescence and classified as negative (102/137) or positive (35/137) after conventional reading by an experienced biologist. Digitized images of microscopic fields were processed with a computerized algorithm yielding 4 quantitative parameters. The obtained dataset was then randomly split 100 times between a training set (102 images) and a test set (35 images). The classification efficiency of three standard classifiers was then assayed on the train and test sets after training on the train set. Mean results of accuracy indices are shown +/- standard error of the mean.

| Analytic tool | Data set | prediction accuracy<br>(pa) | corrected prediction accuracy (cpa) | area under roc curve<br>(auc) |
| --- | --- | --- | --- | --- |
| Logistic Regression | train | 0.92 +/- 0.002 | 0.68 +/- 0.006 | 0.95 +/- 0.001 |
|  | test | 0.91 +/- 0.004 | 0.64 +/- 0.014 | 0.95 +/- 0.004 |
| Nearest Neighbors<br>(3 neighbors) | train | 0.93 +/- 0.002 | 0.73 +/- 0.007 | 0.88 +/- 0.003 |
|  | test | 0.89 +/- 0.005 | 0.56 +/- 0.015 | 0.81 +/- 0.007 |
| Decision Tree<br>(maximum depth: 3) | train | 0.96 +/- 0.002 | 0.83 +/- 0.006 | 0.94 +/- 0.003 |
|  | test | 0.89 +/- 0.005 | 0.56 +/- 0.016 | 0.84 +/- 0.007 |

**Table 2.** Discrimination between cytoplasmic and nuclear patterns by ML processing of 4 indices. 35 Anca-positive sera were concluded to yield a cytoplasmic (9/35) or perinuclear/nuclear (26/35) pattern after conventional reading by an experienced biologist. Digitized images of microscopic fields were processed with a computerized algorithm yielding 4 quantitative parameters. The obtained dataset was then randomly split 100 times between a training set (26/35) and a test set (9/35) . The classification efficienty of three standard classifiers was then assayed. Mean results of accuracy indices are shown +/- standard error of the mean.

| Analytic tool | Data set | prediction accuracy | Corrected accuracy | area under roc curve (auc) |
| --- | --- | --- | --- | --- |
| Logistic Regression | train | 0.84 +/- 0.006 | 0.32 +/- 0.024 | 0.79 +/- 0.006 |
|  | test | 0.77 +/- 0.013 | 0.17 +/- 0.030 | 0.78 +/- 0.030 |
| Nearest Neighbors (3 neighbors) | train | 0.85 +/- 0.004 | 0.39 +/- 0.016 | 0.71 +/- 0.008 |
|  | test | 0.79 +/- 0.011 | 0.23 +/- 0.028 | 0.66 +/- 0.015 |
| Decision Tree (maximum depth: 3) | train | 0.94 +/- 0.004 | 0.73 +/- 0.016 | 0.91 +/- 0.006 |
|  | test | 0.68 +/- 0.014 | 0.04 +/- 0.018 | 0.57 +/- 0.017 |

While classification efficiency might be considered as fairly good as compared to other studies, efficiency parameters were significantly lower than one and it was important to explore different possibilities of improving this situation.

i. When we tried a simple neural network (multilayer perceptron) as a more elaborate model, classification efficiency was not improved (test *cpa*=0.59, test *auc*=0.83), in accordance with our earlier conclusion that simple ML models were better suited to process limited datasets [28].
ii. Since ML is considered as fairly “data hungry”[35], it was of interest to ask whether an insufficient dataset size (137 samples) might be an important cause of prediction errors. This question was addressed by measuring the LR classification efficiency on a series of randomly built subsets. As shown on Figure 3, *index-based* classification efficiency was only weakly dependent on the dataset size.
iii. The behavior of ML algorithms is dependent on so-called hyperparameters that are often ignored since default values are usually satisfactory. It was checked that the classification efficiency of LR could not be improved by changing LR regularization parameter C (not shown) . As expected, the default value (C=1) was found satisfying. Reducing regularization resulted in significant increase of training *cpa*, with a decrease of test *cpa*, which was indicative of overfitting. Increasing regularization resulted in concomitant decrease of *cpa* on training and test datasets (not shown).
iv. Aforementioned results strongly suggested that classification efficiency was limited by intrinsic capacity of indices used to quantify images. Since the the first index was derived from our experience of automatic detection of anti-nuclear antibodies [25] [14], we tested the discrimination provided by this sole index, based on empirical determination of a threshold value separating positive from negative samples. Our dataset was randomly split 100 times between and training set (102 samples) and a test set (35 samples). The average *cpa* parameters obtained on train and test sets were respectively 0.705 +/- 0.004 SE and 0.701 +/- 0.013 SE which were slightly but significantly higher than efficiency parameters shown on Table 1.

**Figure 3.**
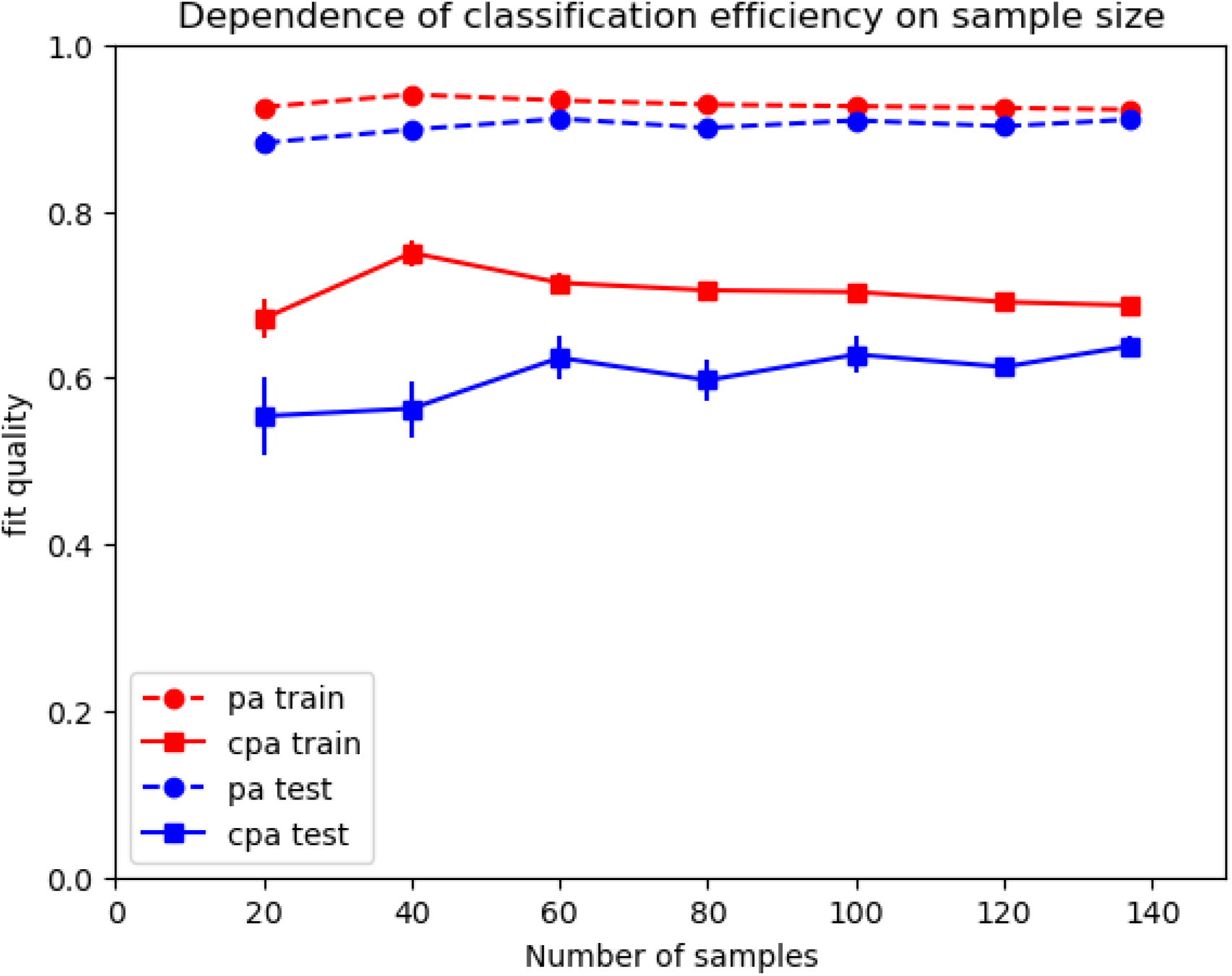
Dependence of index-based classification efficiency on dataset size. A dataset of 137 serum samples was randomly reduced to 20, 40, 60, 80, 100 or 120 samples and the efficiency of positive/negative discrimination by LR was calculated. This process was repeated 100 times for each data size and mean cpa is shown. Vertical bar length is twice the SEM.

#### 3.1.2 Automatic discrimination between several fluorescence patterns

It was felt of interest to determine whether ML could help us discriminate between different fluorescence patterns. We investigated the possibility of automatic discrimination between the 9 C- ANCA positive and 26 P-ANCA or ANA positive images yielded by the 35 positive sera included in our dataset. Interestingly, a preliminary study revealed that none of the four aforementioned indices individually allowed any discrimination between both groups : indeed, the efficiency parameter *cpa* obtained by separating both groups was respectively 0.0447 +/- 0.1530 SD, 0.0545 +/- 0.2101 SD, 0.0300 +/- 0.2127 SD and 0.0252+/- 0.1542 SD with indices i1 to i4. However, when the four-parameter dataset was processed with three ML algorithms, a poor but significant discrimination between C-ANCA and P-ANCA was obtained. Results are displayed on Table2.

Finally, ML was used to process the whole 137 sample dataset in order to try and discriminate between four groups of interest: negative (102/137), C-ANCA (9/137), P-ANCA (21/137) or atypical patterns due to ANAs (5/137). As shown on Table 3, a significant discrimination was obtained.

**Table 3.** Discrimination between four fluorescence patterns by processing a 4-parameter dataset. 137 sera were assayed for Ancas with indirect immunofluorescence and categorized as negative (102/137), C-ANCA (9/137), P-ANCA (21/137) or anti-nuclear (5/137) after conventional reading by an experienced biologist. Digitized images of microscopic fields were processed with a computerized algorithm yielding 4 quantitative parameters. The obtained dataset was then randomly split 100 times between a training set (102 images) and a test set (35 images). The classification efficiency of three standard classifiers was then assayed on the train and test sets after training on the train set. Mean results of accuracy indices are shown +/- standard error of the mean.

| Analytic tool | Data set | prediction accuracy (pa) | corrected prediction accuracy (cpa) |
| --- | --- | --- | --- |
| Logistic Regression | train | 0.87 +/- 0.002 | 0.66 +/- 0.005 |
|  | test | 0.82 +/- 0.005 | 0.61 +/- 0.012 |
| Nearest Neighbors (3 neighbors) | train | 0.90 +/- 0.002 | 0.75 +/- 0.006 |
|  | test | 0.81 +/- 0.006 | 0.56 +/- 0.014 |
| Decision Tree (maximum depth: 3) | train | 0.90 +/- 0.002 | 0.80 +/- 0.005 |
|  | test | 0.79 +/- 0.007 | 0.57 +/- 0.013 |

The relatively small difference between performance parameters obtained with three different algorithms and impossibility to improve agreement with hyperparameter adaptation suggested that the limitation was essentially due to an insufficient discriminative power of the four-feature description used to account for image properties. However, these results were consistent with the widespread hypothesis that a simple strategy for achieving automatic classification of ANCA-related images might consist of adding an increasing number of texture parameters and automatically combining them with simple ML algorithms. Indeed, ANA pattern analysis was performed with a commercial system involving 1400 object-describing parameters [36]. However, recent progress of AI was an incentive to look for a fully autonomous way of analysing immunofluorescence images. Results obtained along this line are show below.

### 3.2 Use of AI for autonomous analysis of fluorescence images

Two strategies were considered : i) combining data reduction with fairly simple ML algorithms. ii) using neural networks for complete analysis.

#### 3.2.1 Use of data reduction to process invidual cell images

The description of analyzed microscope fields with only four global parameters was replaced with the use of a 2,500 parameter set (50×50 pixel intensities) to account for each cell image contained in a given microscope field. Fifty one sera were used to build a dataset of 1,733 individual cell images (513 negative, 309 C-ANCA, 789 P-ANCA and 122 atypical patterns that could be ascribed to ANAs).

In a first step, the capacity of aforementioned three standard ML algorithms to analyse these images without any data reduction was studied. As shown on Table 4, parameter *cpa* obtained for positive/negative discrimination was fairly low. Since the important difference between train and test *cpa* was indicative of overfitting that might be ascribed to an excessive number of features as compared to the sample number, we used principal component analysis as a standard way of reducing the number of parameters. As shown in Table 4, this resulted in significant increase of efficiency parameters since the highest test *auc* was raised from 0.86 to 0.92 and the highest test *cpa* was increased from 0.35 to 0.46.

**Table 4.**
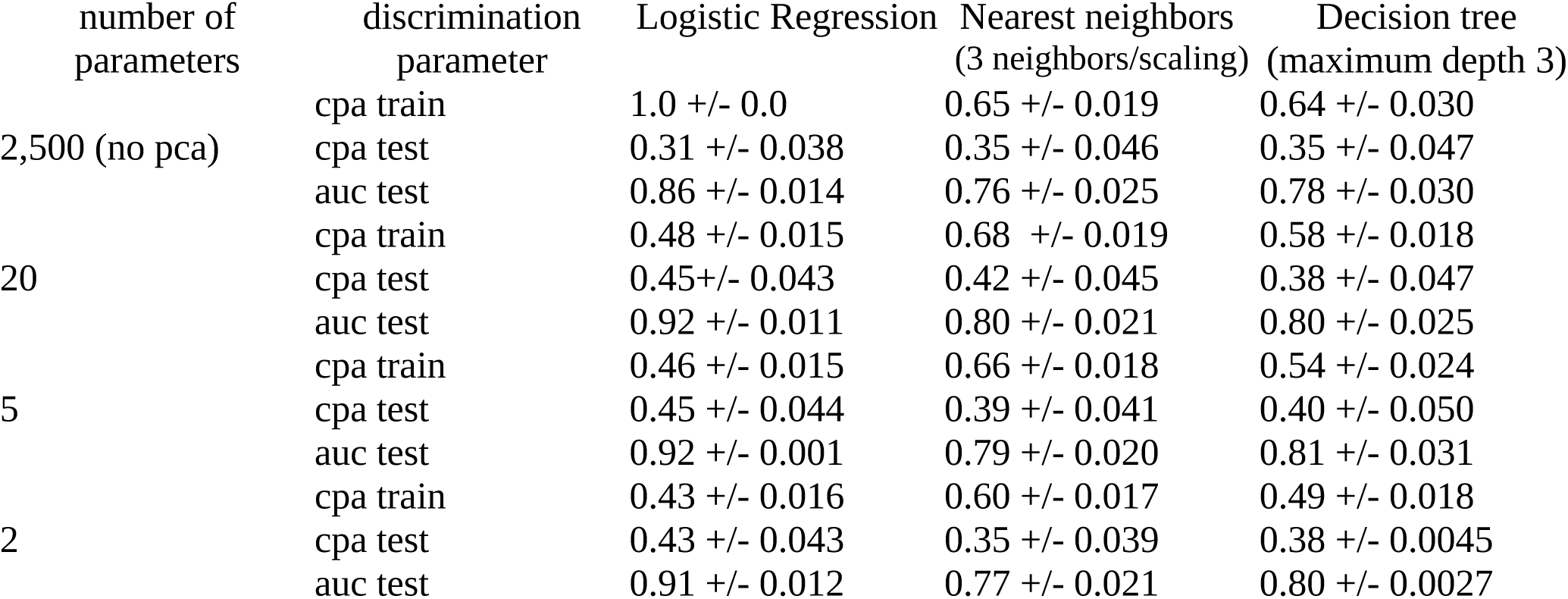
Discrimination between Anca-positive and negative sera by processing 2,500 pixel images. 1733 images from 51 microscope fields were assayed for ANCAs with indirect immunofluorescence and categorized as negative (513/1733) or positive (1220/1733) after conventional reading by an experienced biologist. Digitized images of microscopic fields (50×50 pixel were analyzed as 2,500 parameter objects or preprocessed with principal component analysis for retaining the main 20, 5 or 2 components. Datasets were then randomly split 100 times between a training set (1299 images) and a test set (434 images). Three simple algorithms were then trained and assayed for discriminative efficiency. Mean values of corrected predictive accuracy (*cpa*) for train and test datasets, and area under roc curve (*auc*) for test dataset are shown +/- standard deviation

| number of<br>parameters | discrimination<br>parameter | Logistic Regression | Nearest neighbors<br>(3 neighbors/scaling) | Decision tree<br>(maximum depth 3) |
| --- | --- | --- | --- | --- |
| 2,500 (no pca) | cpa train | 1.0 +/- 0.0 | 0.65 +/- 0.019 | 0.64 +/- 0.030 |
|  | cpa test | 0.31 +/- 0.038 | 0.35 +/- 0.046 | 0.35 +/- 0.047 |
|  | auc test | 0.86 +/- 0.014 | 0.76 +/- 0.025 | 0.78 +/- 0.030 |
| 20 | cpa train | 0.48 +/- 0.015 | 0.68 +/- 0.019 | 0.58 +/- 0.018 |
|  | cpa test | 0.45 +/- 0.043 | 0.42 +/- 0.045 | 0.38 +/- 0.047 |
|  | auc test | 0.92 +/- 0.011 | 0.80 +/- 0.021 | 0.80 +/- 0.025 |
| 5 | cpa train | 0.46 +/- 0.015 | 0.66 +/- 0.018 | 0.54 +/- 0.024 |
|  | cpa test | 0.45 +/- 0.044 | 0.39 +/- 0.041 | 0.40 +/- 0.050 |
|  | auc test | 0.92 +/- 0.001 | 0.79 +/- 0.020 | 0.81 +/- 0.031 |
| 2 | cpa train | 0.43 +/- 0.016 | 0.60 +/- 0.017 | 0.49 +/- 0.018 |
|  | cpa test | 0.43 +/- 0.043 | 0.35 +/- 0.039 | 0.38 +/- 0.0045 |
|  | auc test | 0.91 +/- 0.012 | 0.77 +/- 0.021 | 0.80 +/- 0.0027 |

The possibility of discriminating beween nuclear/perinuclear and cytoplasmic fluorescence was also studied. As shown on Table 5, standard algorithms displayed a poor capacity to discriminate between both patterns, since the maximum *cpa* value was about 0.10, when determined on full images or on the first 20 principal components.

**Table 5.** Discriminating between cytoplasmic and nuclear patterns by processing whole cell images. 1220 cell images classified as C-ANCA (309/1220) or with a nuclear/perinuclear pattern (911/1220) after conventional reading by an experienced biologist were processed with three standard ML algorithms. The obtained dataset was then randomly split 100 times between a training set (915/1220) and a test set (305/1220) . The classification efficiency of three standard classifiers was then calculated either on full sets of pixel intensities (2,500 pixels per image) or using the first 20 components yielded by principal component analysis. Mean results of accuracy indices are shown +/- Standard Deviation.

| ML algorithm | Data set | prediction accuracy<br>(pa) | Corrected accuracy<br>(cpa) | area under roc curve<br>(auc) |
| --- | --- | --- | --- | --- |
| Logistic Regression | train full | 1.00 +/- 0.00 SD | 1.00 +/- 0.00 SD | 1.00 +/- 0.00 SD |
|  | test full | 0.67 +/- 0.024 SD | 0.04 +/- 0.022 SD | 0.54 +/- 0.019 SD |
|  | train 20c | 0.76 +/- 0.008 SD | 0.06 +/- 0.018 SD | 0.53 +/- 0.009 SD |
|  | test 20c | 0.75 +/- 0.020 SD | 0.03 +/- 0.021 SD | 0.52 +/- 0.011 SD |
| Nearest Neighbors<br>(3 neighbors,scaling) | train full | 0.84 +/- 0.006 SD | 0.40 +/- 0.017 SD | 0.74 +/- 0.011 SD |
|  | test full | 0.71 +/- 0.022 SD | 0.08 +/- 0.034 SD | 0.55 +/- 0.025 SD |
|  | train 20c | 0.85 +/- 0.007 SD | 0.42 +/- 0.023 SD | 0.74 +/- 0.012 SD |
|  | test 20c | 0.73 +/- 0.019 SD | 0.11 +/- 0.033 SD | 0.58 +/- 0.022 SD |
| Decision Tree<br>(maximum depth: 5) | train full | 0.83 +/- 0.015 SD | 0.34 +/- 0.032 SD | 0.68 +/- 0.33 SD |
|  | test full | 0.73 +/- 0.023 SD | 0.09 +/- 0.038 SD | 0.56 +/- 0.023 SD |
|  | train 20c | 0.81 +/- 0.013 SD | 0.28 +/- 0.048 SD | 0.65 +/- 0.036 SD |
|  | test 20c | 0.55 +/- 0.023 SD | 0.07 +/- 0.036 SD | 0.55 +/- 0.023 SD |

Since images were expected to include the information required to discriminate between C-ANCA and P-ANCA, it was of obvious interest to try and determine why standard ML algorithms were unable to select the relevant information from image. A likely possibility might be that each serum generated a particular fluorescence pattern, in addition to a “general” cellular or nuclear localization. Training would thus result in a capacity of a ML model to recognize patterns specific for the particular antibody set of each tested serum. This possibility was addressed by visualization of the first two principal components of images displayed by six sera (3 cANCA, 3 pANCA), as shown on Figure 4 a clearcut separation could be observed between images generated by different sera of similar (either C-ANCA or P-ANCA) specificity.

**Figure 4.**
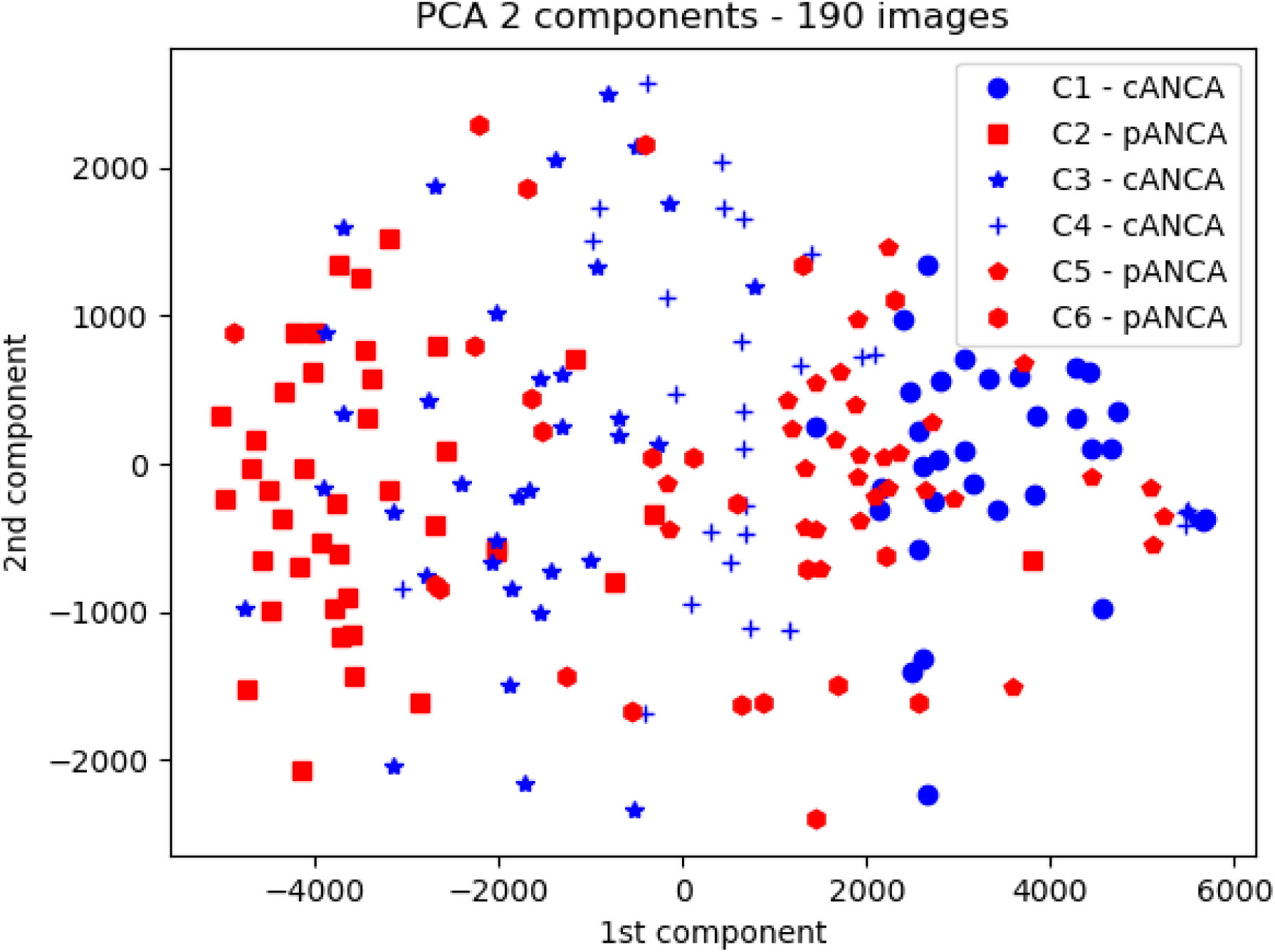
Six microscope images generated with 6 sera (3 C-ANCA, 3 P-ANCA) yielded 190 individual cell images. Pixel intensities were subjected to principal component analysis, and the first two components are displayed. Clearly, C-ANCA (blue) and P-ANCA (red) displayed marked orverlap, but images corresponding to a same serum displayed significant separation.

This supported the need for a more refined ML algorithm allowing precise selection of desired features. In view of the remarkable success obtained with neural networks in the field of image analysis, it was deemed appropriate to use a number of neural-networks to analyze our dataset.

#### 3.2.2 Analysis of full images with neural networks

While recent successes met by neural networks in the field of image analysis were an incentive to explore the potential of this model class, a major problem is that a neural network may involve a high number of hyperparameters. First, we used Mutilayer perceptron as relatively simple models and the importance of three major hyperparameters (hidden layer number, hidden layer size, and regularization parameter) is shown on Fig 5.

**Figure 5.**
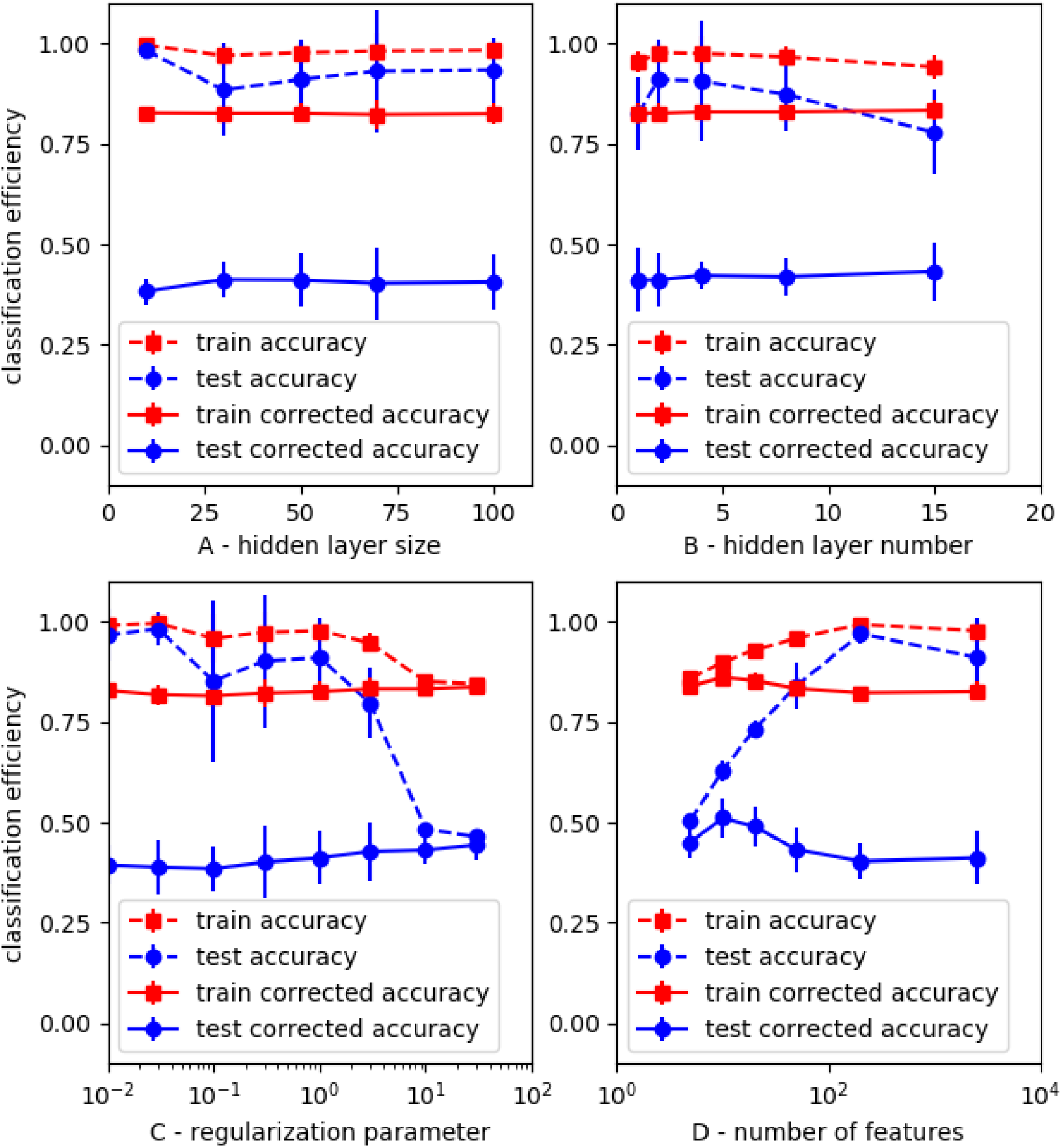
Dependence of Mlp efficiency on model settings. A series of neural networks with varying number and size of hdden layers and regularization parameter were used to discriminate between positive (513) and negative (1200) samples in a set of 1733 images. For each hyperparameter combination, this set was randomly split between 10 and 40 times into a trainin and test set. In another series of calculations, PCA was used for data reduction. Mean values of efficiency parameters are shown. Vertical bar line is twice the standard error.

The following conclusions were suggested :

- i) efficiency parameters displayed limited change in response to fairly extensive variation of hyperparameters, suggesting a moderate dependence of classification efficienty on the choice of model hyperparameters.
- ii) Parameter *cpa* yielded on test sets varied between a minimum value of 0.38 and a maximum of 0.51 (with *kappa score* and *auc* respectively equal to 0.67 and 0.85). Neural network performance was thus better than that achieved with standard ML models (shown on Table 4).
- iii) plots displayed of Figure 5 C&D clearly confirmed the risk of overfitting as a consequence of insufficient regularization (C) or excessive number of features (D) as compared to the number of samples, leading to a high *cpa* train/*cpa* test ratio.

These results were an incentive to investigate the capacity of Mlp to discriminated between different fluorescent patterns. When an dataset of 1220 images with cellular (309) or nuclear/perinuclear (911) fluorescence localization was studied with a wide range of hyperparameter settings, test *cpa* ranged between 0 and 0.11, suggesting that this dataset was insufficient to allow proper model training, as was also found with simpler models (Table 5).

An important point is that high *cpa* values for positive/negative and pattern discrimination of the images of *training sets* were obtained, since the highest values were respectively 0.99 and 0.92. These results suggested the conclusion that ML models were sufficiently versatile to efficiently discriminate between all images, but the classification criteria obtained with autonomous training did not match the biologically significant classification.

A possible reason for the limitation of Mlp efficiency is that the localization of individual pixels might not be sufficiently apparent in parameter sets organized as 1-dimensional arrays. Indeed, convolutional neural networks may be considered as better suited than networks including only so-called dense layers to detect specific image patterns when they are trained with 2-dimensional arrays. Thus, we tentatively assayed the capacity of a series of convolutional neural networks to perform positive/negative classification of 1733 images data sets. Test *cpa* values of 0.52, comparable with those obtained with Mlp, were obtained. Also, no significant improvement of pattern classification was obtained with aforementioned 1200 image dataset.

A possible explanation for neural network limitation might be that differences between individual sera and cells might generate fluorescence variations overlapping the effects of antibody specificity. A simple way of testing this possibility consisted of performing **controlled splitting** of image sets by ensuring that all images generated by a given serum were in the same (training or test) set. Results obtained with this strategy are shown below.

#### 3.2.3 Combination of controlled splitting of train and test datasets and serum rather than image classification

The following two modifications of processing were performed: (i) it was ensured that all images generated by a same serum fell into the same (train or test dataset); (ii) after training models on 2500 pixel images as indicated above, sera belonging to test datasets were classified as positive or negative according to the highest number of individual cell images classified as positive or negative.

As shown on Table 6, controlled splitting did not improve individual cell classification, but the modified procedure resulted in highly significant improvement of serum classification, since test *cpa* was 0.74 with Mlp (the corresponding kappa score was 0.84).

**Table 6.**
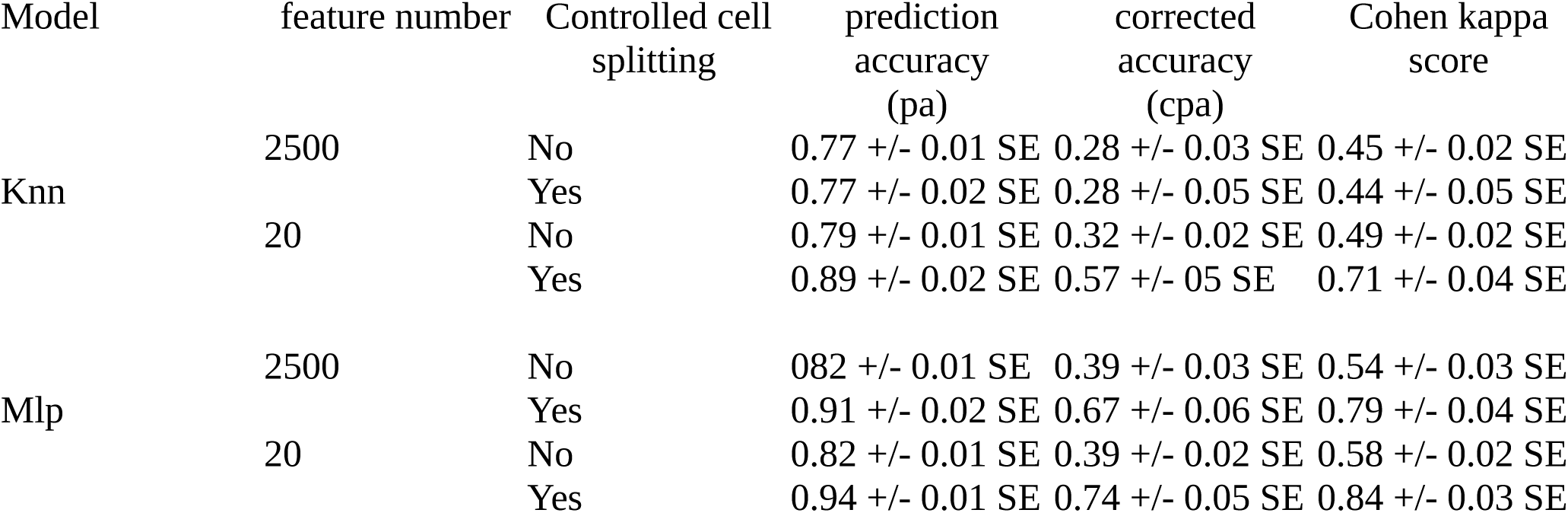
Discrimination between ANCA-positive and negative sera by processing of individual cell images. Fifty-one (35 positive, 16 negative) sera were tested for ANCA detection with immunofluorescence: 1,733 cell images were processed with two machine-learning algorithms: k-nearest neighbors (Knn, n=3 neigbors) and multilayer perceptron (Mlp, one 40-neuron hidden layer) for positive/negative discrimination. The image dataset was randomly split either without any restriction (no control) or with ensuring that all images generated with a given sera were gathered in the same (training or test) group. This process was repeated 25 fold and mean values of efficiency parameters are displayed together with standard error of the mean (SE). The prediction accuracy was calculated either for each cell (no control group) or for each serum, by classifying each serum as the most frequent classification of corresponding cell images.

| Model | feature number | Controlled cell splitting | prediction accuracy (pa) | corrected accuracy (cpa) | Cohen kappa score |
| --- | --- | --- | --- | --- | --- |
| Knn | 2500 | No | 0.77 +/- 0.01 SE | 0.28 +/- 0.03 SE | 0.45 +/- 0.02 SE |
|  |  | Yes | 0.77 +/- 0.02 SE | 0.28 +/- 0.05 SE | 0.44 +/- 0.05 SE |
|  | 20 | No | 0.79 +/- 0.01 SE | 0.32 +/- 0.02 SE | 0.49 +/- 0.02 SE |
|  |  | Yes | 0.89 +/- 0.02 SE | 0.57 +/- 0.05 SE | 0.71 +/- 0.04 SE |
| Mlp | 2500 | No | 0.82 +/- 0.01 SE | 0.39 +/- 0.03 SE | 0.54 +/- 0.03 SE |
|  |  | Yes | 0.91 +/- 0.02 SE | 0.67 +/- 0.06 SE | 0.79 +/- 0.04 SE |
|  | 20 | No | 0.82 +/- 0.01 SE | 0.39 +/- 0.02 SE | 0.58 +/- 0.02 SE |
|  |  | Yes | 0.94 +/- 0.01 SE | 0.74 +/- 0.05 SE | 0.84 +/- 0.03 SE |

It was investigated whether this controlled splitting might improve pattern classification. However, classification efficiency was not improved - and was in fact significantly decreased - when this controlled splitting was performed (not shown).

## 4 Discussion

The main purpose of this work was to investigate a fairly simple and well-defined problem of current medical interest to explore the possibility of autonomous building of ML models with sufficient reliability to assist and possibly replace biological experts in analyzing microscopic images. It was hoped that this endeavour might help identifying general guidelines, limitations and possible strategies for future progress.

A first conclusion is that currently available ML algorithms autonomously trained on a fairly restricted dataset were able to perform positive/negative discrimination with substantial or even good agreement with experienced human analysis (highest kappa score was 0.84). This mere finding is of actual clinical interest. Indeed, automatic positive/negative discrimination would be most useful by decreasing current delay is delivering a negative diagnosis or performing Elisa assays.

It must be emphasized that the estimated values of achieved efficiency parameters may be considered as fairly reliable despite the lack of a fully independent validation dataset. Indeed, average values of efficiency indices were estimated after repeated - up to 100 fold - splitting of the full data set, and results shown on Figure 5 and other tests performed on basic classifiers suggest that these efficiency parameters were not strongly dependent on model hyperparameters, thus making less likely the possibility that calculate efficiency indices might be artefactually high due to a strong influence of the dataset on parameter choice. In any case, as was recently emphasized [37] [38], safe model validation would require to perform additional check on microscopical images obtained under fairly independent conditions.

The second conclusion is that our simple approach did not succeed in discriminating between different fluorescence patterns. Two non exclusive strategies may be considered to achieve further progress: i) using more sophisticated algorithms to match human experience, as was done on ANA research [11] or histopathologic studies [39], which would require to prepare a substantially larger databas, or ii) by combining different fixation or staining procedures, to facilitate image analysis, e.g. by analyzing both ethanol- and paraformaldehyde-fixed cells [8] . Indeed, C-ANCA might thus be more easily recognized on a basis of a different fluorescence localization yielded by different fixation procedures .

A third conclusion is that simple models, such as kNN may be more rewarding that complex neural networks to perform simple classification with limited datasets. Indeed, model settings are easier to select with simple models, also, results obtained with conceptually simple models may be more easily interpreted that those yielded by models as complex as neural networks that may involve more than one million parameters [40], which might facilitated further progress.

As shown in the first part of this report (Table 1), the use of biologically inspired indices is an attractive way of combining biological expertise and AI. Indeed, many commercially available systems were based on the use of ML to process extensive sets of texture parameters. However, the development and continuous improvement of an algorithm involving more than 1,000 parameters [36] may be more difficult to perform than the autonomous building of ML models with general strategies that did not rely on any specific expertise to determine model architecture.

A general conclusion of our study is that an essential requirement for further progress will consist of increasing the size of our database. It would also be useful to review the individual classification errors concerning cell images in order to improve our understanding of the origin of model limitation. Such an increased database might allow us to build a reliable classification model yielding at the same time positive/negative diagnosis and discrimination between different fluorescence patterns that might be indicative of anti-myeloperoxidas, anti-proteinase 3, AN, or hopefully of new antibody specificities indicative of other pathological situations requiring different therapeutic management, such as acute infection.

## Data Availability

All data produced in the present will be made available after proper agreement from the institution ethics committee

## Abbreviations

AI: artificial intelligence
ANA: anti nuclear antibody
ANCA: anti-neutrophil cytoplasmic antibody
auc: area under roc curve
C-ANCA: cytoplasmic type ANCA
cpa: corrected prediction accuracy
DT: decision tree
kNN: k nearest neighbors
LR: logistic regression (classifier)
ML: machine learning
Mlp: multilayer perceptron
P-ANCA: perinuclear-type
ANCA pa: prediction accuracy
ROC: receiver operator curve

## Acknowledgments

All specific apparatus used in this study was funded by the grant “AORC Junior 2017” from the Assistance Publique Hôpitaux de Marseille (APHM, Marseillle France)

## Author contribution

DB, PB and NB devised the study. DB and NB managed biological investigations. PB performed IA calculations and drafted the manuscript. DB, PB and NB finalized the manuscript.

